# OCPD Symptoms in Veterans Receiving PTSD Specialty Care

**DOI:** 10.64898/2026.06.24.26356458

**Authors:** Jennifer Barredo, Meghan J. Kulak, Hannah R. Swearingen, Timothy Y. Mariano, M. Tracie Shea, Anthony Pinto, Benjamin D. Greenberg

## Abstract

Post-traumatic stress disorder (PTSD) is associated with high rates of comorbid personality disorders, which may contribute to PTSD severity. Among veterans with PTSD, obsessive compulsive personality disorder (OCPD) is common, with reported prevalence estimates ranging from 7-44%. Despite this, the relationship between OCPD traits and PTSD severity remains poorly understood. This retrospective, cross-sectional study examined associations between PTSD severity and OCPD traits in a naturalistic sample of 99 Veterans evaluated by a single clinician in a PTSD/Trauma Recovery Services clinic. PTSD symptoms were measured with the PTSD Checklist for DSM-V (PCL-5), and OCPD traits were measured with the Pathological Obsessive-Compulsive Personality Scale (POPS). Relationships between these two constructs were examined using Pearson’s correlations. Overall PTSD severity was significantly and positively correlated with total OCPD traits (*r* = 0.46, *p* < 0.001). Among OCPD domains, maladaptive perfectionism showed the strongest association with PTSD severity (*r* = 0.44, *p* <.001), followed by emotional overcontrol and reluctance to delegate (both *r* = .38, *p* < .01), rigidity (r = .35, p < .01), and difficulty with change (r = .28, p < .05). These findings suggest OCPD traits impact PTSD symptom burden in veterans, warranting further research and clinical attention.

**Public Significance:** Veterans living with post-traumatic stress disorder (PTSD) have high rates of obsessive-compulsive personality disorder (OCPD) traits such as rigidity, perfectionism, and emotional overcontrol. This study found that in one VA clinic, veterans with higher levels of OCPD traits tended to experience more severe PTSD symptoms. Understanding how personality traits impact trauma recovery may help clinicians tailor treatments more effectively.

## Introduction

Posttraumatic stress disorder (PTSD) is a leading contributor to functional impairment and suicide risk among veterans (Rassu et al., 2022). Yet even within trauma-exposed populations, PTSD varies widely in symptom severity, functional impact, and treatment response, and the sources of this heterogeneity remain incompletely understood (Campbell-Sills et al., 2022; Grau et al., 2022). Personality factors may be one important, but underexamined contributor. In particular, obsessive-compulsive personality disorder (OCPD), characterized by a pervasive need for perfection and control that constrains functioning and quality of life, may influence the emergence and persistence of trauma-related symptoms.

OCPD can be difficult to recognize because cardinal features like conscientiousness, discipline, high standards, and attention to detail may appear adaptive, especially in military and veteran contexts. What distinguishes OCPD is not the presence of these traits, but their inflexibility, extremity, and cost. Core features include maladaptive perfectionism, emotional overcontrol, rigidity, and rule-bound behavior (*Diagnostic and Statistical Manual of Mental Disorders*, 2022) These traits confer substantial psychiatric vulnerability, reflected in high rates of comorbidity (Grant et al., 2012). Because OCPD traits are often experienced as ego-syntonic (Pinto et al., 2022), patients may not identify them as concerns. Moreover, clinicians may overlook OCPD when more familiar conditions such as PTSD, depression, or anxiety disorders are present. As a result, OCPD may be more likely to go undiagnosed and untreated, with unaddressed maladaptive patterns complicating co-occurring clinical presentations, as well as treatment response (Pinto et al., 2022).

The clinical relevance of OCPD to PTSD is compelling because hallmarks of overcontrolled personality functioning align with processes implicated in the maintenance of trauma-related distress, such as emotional regulation problems (McLean & Foa, 2017), experiential avoidance (Gerdan & Salcioglu, 2025; Kelly et al., 2019; Patel et al., 2023), and interpersonal strain (Beck et al., 2009). For example, the strong desire for control may limit disclosure, emotional awareness, and interpersonal connection with therapists and other supportive individuals. Maladaptive perfectionism may intensify trauma-related shame and self-criticism. Rigidity may restrict openness to learning new strategies for coping with trauma reminders, undermining treatment engagement. When OCPD traits co-occur with PTSD, their dynamics may both maintain and intensify PTSD symptom burden.

OCPD is one of the most common personality disorders in the general population, with an estimated global prevalence rate of 6.5% (Clemente et al., 2022). It appears to be overrepresented among veterans, with prevalence estimates ranging from 17–44% (Bollinger et al., 2000; Dunn et al., 2004; Southwick et al., 1993). Despite this, OCPD remains underdiagnosed in clinical settings. In veteran healthcare systems, this underrecognition may be reinforced by diagnostic and structural disincentives(Edwards et al., 2022). Hence, overcontrolled personality traits may be common among trauma-exposed Veterans, but their contribution to PTSD severity and treatment complexity remains poorly characterized.

The present naturalistic chart review extends the limited literature on OCPD in Veteran populations by examining the prevalence and clinical correlates of OCPD traits among Veterans receiving trauma-related outpatient mental health care within a single psychiatrist’s clinic. We evaluated whether greater OCPD trait burden was associated with greater PTSD severity. By characterizing these associations in a real-world clinical setting, this study aims to clarify whether overcontrolled personality pathology contributes to clinical complexity in trauma-focused Veteran care and to identify whether systematic attention to OCPD traits may improve case formulation and treatment planning.

## Methods

The primary aim of this chart review was to examine associations among PTSD symptom severity and obsessive-compulsive personality traits. The local Institutional Review Board (IRB) approved this chart review as an exempt study for secondary data use. A waiver of informed consent was granted by the

IRB, due to the minimal risk nature of the study and impracticability of obtaining consent. A Health Insurance Portability and Accountability Act (HIPAA) waiver was also granted permitting retrospective review of patient assessments collected during the course of clinical practice and treatment as usual.

### Setting and Data Source

Clinical data were obtained from medical records of veterans seen for an initial psychiatric consult with one author (BDG), a staff psychiatrist in the Trauma Recovery Services clinic at the Providence VA Medical Center between January 2016 and August 2025. Veterans were eligible for inclusion if their initial consultation paperwork contained a completed Pathological Obsessive-Compulsive Personality Scale (POPS). Clinic patients completed the POPS as a course of routine care during their initial consultation with the author. PTSD Checklist (PCL) scores were only available for a subsample of patients, as PTSD was documented in some patients referred to this clinic using alternative measures (e.g., Clinician-Administered PTSD Scale for *DSM-5*). Clinical records were reviewed to obtain demographic information for all eligible veterans.

### Clinical Measures

#### Pathological Obsessive-Compulsive Personality Scale (POPS)

The POPS (Pinto et al., 2011) is a 49-item self-report measure of obsessive-compulsive personality traits and severity that includes five specific trait factors and an overall factor (total score), assessing obsessive-compulsive personality pathology severity and dysfunction. The total and subscale scores have demonstrated strong internal consistency and convergent and discriminant validity with other measures of personality disorders (Sadri et al., 2019).

#### PTSD Symptom Checklist for DSM-5 (PCL-5)

The PCL-5(F. Weathers, 2013) is a widely used self-report measure of PTSD. It includes 20 items that correspond to the 20 PTSD symptoms in the DSM-5, with a total score ranging from 0 to 80. PCL-5 scores have shown good internal consistency, test-retest reliability, and convergent and discriminant validity (F. W. Weathers et al., 2018).

### Statistical Analysis

Python 3 with compatible pandas, numpy, scipy, and matplotlib packages were used for analyses and visualizations. Descriptive statistics (means, standard deviations, and score distributions) were calculated to characterize PTSD symptom severity (PCL-5) and obsessive-compulsive personality traits (POPS). We visually inspected score distributions using histograms and scatterplots to assess central tendency and distributional shape, with Shapiro-Wilk tests used to evaluate normality. Unless otherwise specified, statistical analyses were conducted using Pearson’s correlations. All statistical tests were two-tailed with α set at .05. Primary analyses examined the relationship between POPS total scores and PCL total scores among veterans with available data. Secondary analyses examined associations between PCL scores and each POPS subscale (Rigidity, Emotional Overcontrol, Maladaptive Perfectionism, Reluctance to Delegate, and Difficulty with Change).

## Results

### Sample Characteristics

The chart review identified 99 veterans (90.9% male) with POPS data available from their initial consultation. Of these, 68 veterans had PTSD symptom data (PCL-5). The mean age was 47.1 years old in the full sample (range 19 to 77 years), and 50.4 years old in the smaller subsample with PCL scores. Other demographics characteristics are summarized in Table 1.

**Table 1.** Race and ethnicity.

|  | White | Black or<br>African<br>American | Asian or<br>Pacific<br>Islander | American<br>Indian or<br>Alaskan Native | Multi-Racial | Other | Unknown | Total |
| --- | --- | --- | --- | --- | --- | --- | --- | --- |
| Hispanic | 8 | 2 | 0 | 0 | 0 | 0 | 0 | 10 |
| Non-Hispanic | 77 | 5 | 4 | 0 | 1 | 0 | 1 | 88 |
| Unknown | 0 | 0 | 0 | 0 | 0 | 0 | 1 | 1 |
| Total | 85 | 7 | 4 | 0 | 1 | 0 | 2 | 99 |

### OCPD Trait Prevalence

The mean POPS total score was 175.78 (SD = 37.57), suggesting that OCPD symptoms were clinically meaningful within this sample, approaching the 80% sensitivity threshold reported in a study examining comorbidity in individuals with OCD in a specialized outpatient clinic (total POPS > 177; (Teller et al., 2026)). Indeed, 55 veterans met the threshold for clinically significant OCPD (total POPS > 177; 55.6%).

### PTSD Symptom Severity and OCPD Traits

Among the 68 veterans with both POPS and PCL scores, 52.9% had an overall POPS score meeting or exceeding the clinically meaningful threshold >177 (n=36). In this subsample, the mean PCL total score was *M* = 46.65 (*SD* = 18.61), indicating overall moderate to high PTSD symptom severity (Figure 1). We observed a moderate, positive correlation between PTSD symptom severity and OCPD traits, *r* = .48, *p* < .001, *n* = 68 (Figure 2).

**Figure 1.**
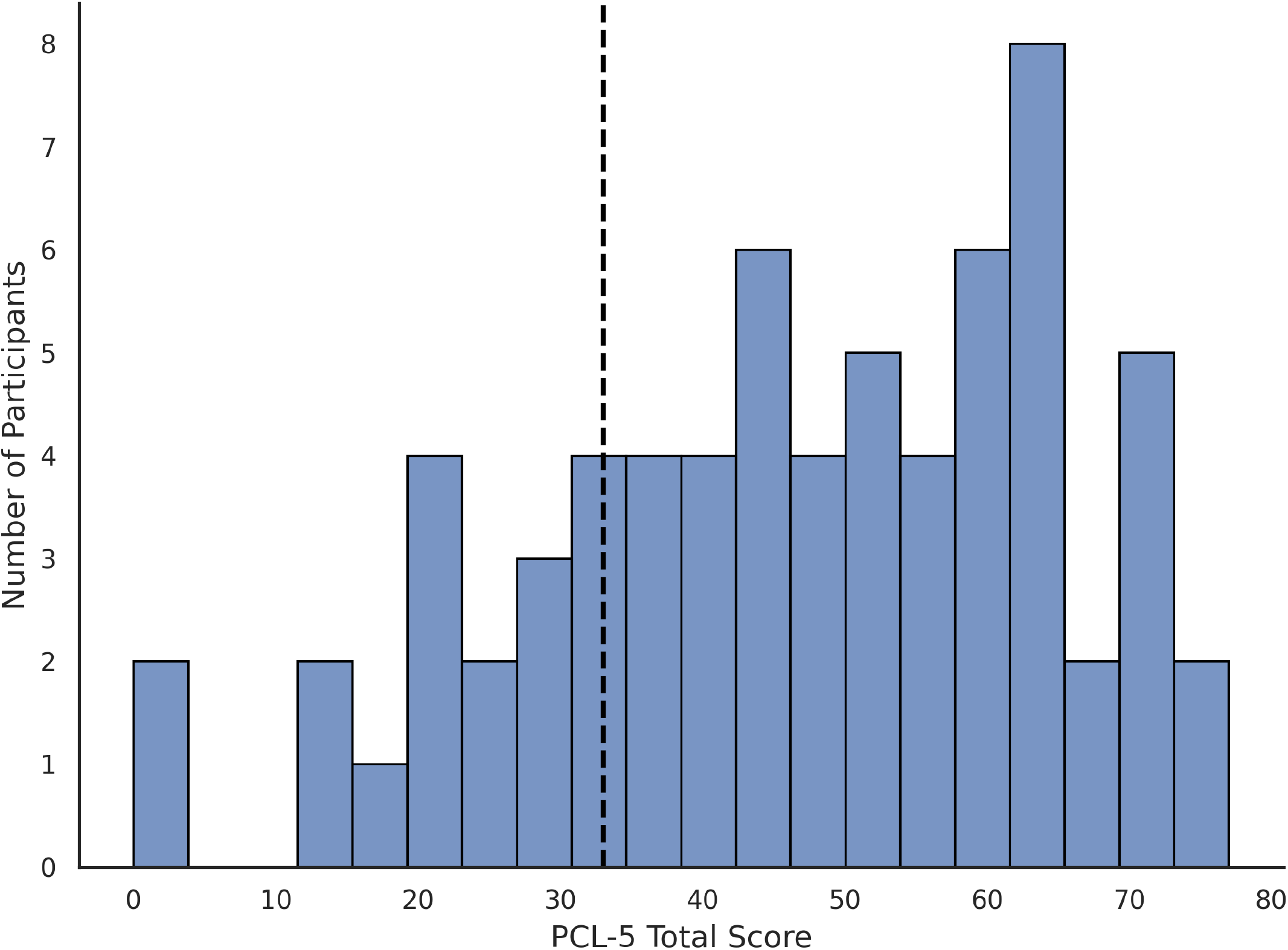
Distribution of PCL-5 total scores for 68 veterans. The dashed line corresponds to an overall score of 33, indicative of clinically significant severity and often considered suggestive of probable PTSD diagnosis.

**Figure 2.**
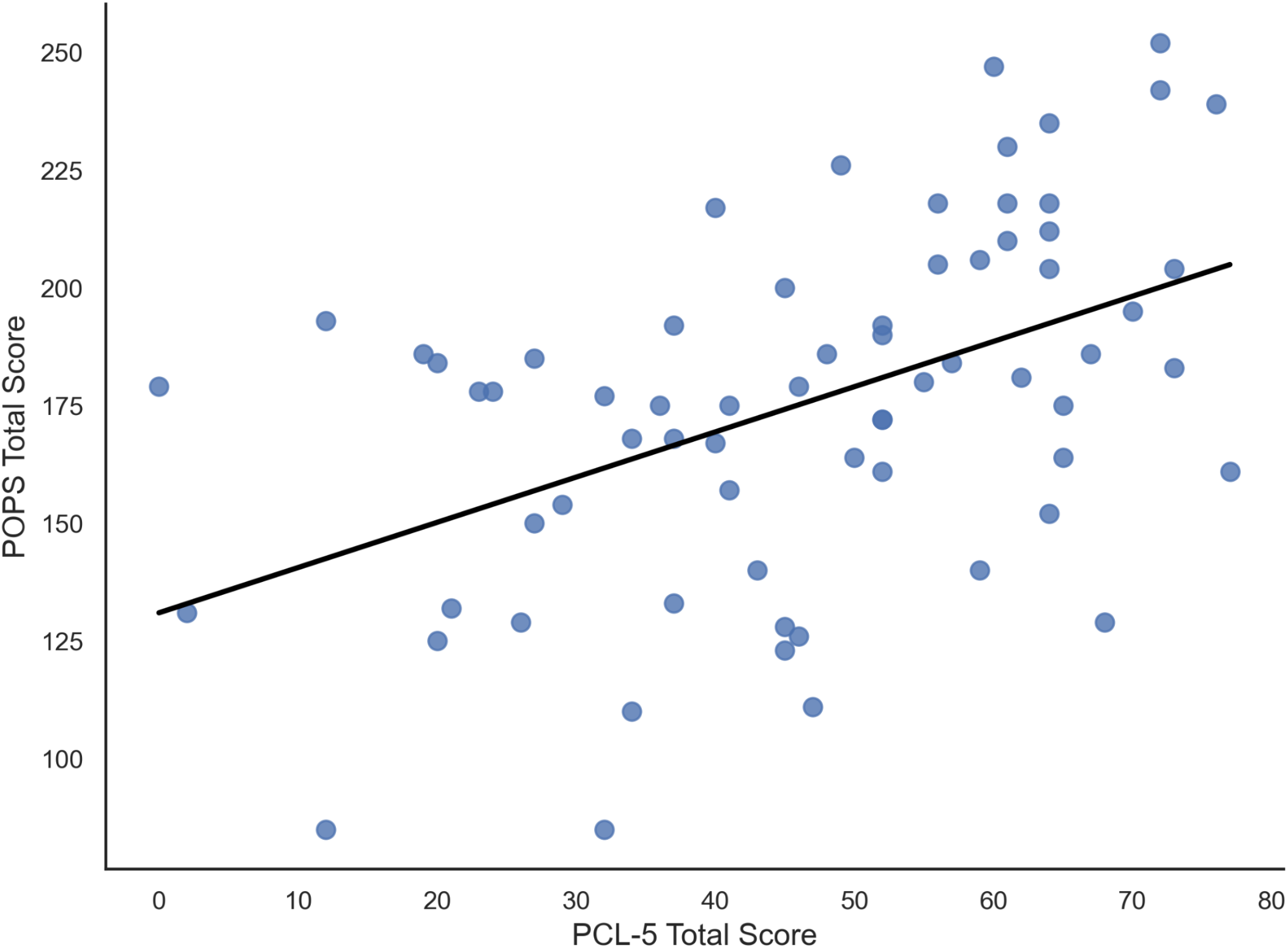
Scatterplot illustrating the relationship between PCL-5 total scores and POPS total scores. The solid line represents the fitted linear regression (Pearson *r* = .48, *p* < .001).

### Associations With OCPD Subscales

PTSD severity was positively associated with all five OCPD domains. The strongest association was observed for maladaptive perfectionism, *r* = .44, *p* < .001, followed by emotional overcontrol, *r* = .38, *p* < .01, and reluctance to delegate, *r* = .38, *p* < .01, then rigidity, *r* = .35, *p* < .01. A smaller, but still significant association was observed for difficulty with change, *r* = .28, *p* < .05.

## Discussion

In this naturalistic chart review study of outpatient veterans, we demonstrate that OCPD traits were significantly associated with PTSD symptom severity. Among the POPS subscales, emotional overcontrol and maladaptive perfectionism showed the strongest associations with PTSD severity. These findings suggest that OCPD, while under-considered in PTSD research or clinical practice, may be a contributor to symptom burden among veterans diagnosed with PTSD in VA care.

These findings are meaningful because OCPD often goes undetected. Despite its high prevalence, it remains underdiagnosed, and this may be especially true in veteran populations. Personality disorder diagnoses have historically carried unfavorable implications for service-connected disability benefits. Clinicians may be reluctant to assign a diagnosis that can be perceived as characterological, and veterans may minimize symptoms that may reflect personality rather than trauma.

But underdiagnosis is not only a documentation problem, it is also a recognition problem. The clinical presentation of OCPD can be subtle, heterogeneous, and difficult to parse from aspects of PTSD. Prior work supports a two-factor model of OCPD comprising a controlling subtype and an anxious subtype (Ansell et al., 2010; Pinto et al., 2022; Riddle et al., 2016). The controlling subtype is more readily recognizable, characterized by rigid rules and routines, verbal hostility, anger outbursts, and efforts to impose high perfectionistic standards on oneself and others. Clinically, this may present as inflexibility, irritability, interpersonal conflict, patterns that generate a high degree of frustration in both the individual and those around them. In contrast, the anxious subtype may be harder to detect. Rather than presenting as domineering or hostile, these individuals often appear conscientious, conflict avoidant, self-critical, excessively detail-focused, and prone to procrastination or difficulty completing tasks. Many patients show features of both, with different traits becoming more visible depending on context (Pinto et al., 2022).

This heterogeneity has important implications for veterans with PTSD. While both presentations may be maladaptive, they interfere with functioning and treatment in different ways. When controlling aspects predominate, anger, interpersonal conflict, and emotion regulation difficulties associated with OCPD may become heightened in the context of comorbid PTSD (Pinto et al., 2022). In contrast, the anxious subtype may reinforce avoidance, and functional impairment in PTSD. Notably, the two OCPD features most strongly associated with PTSD severity in this study, emotional overcontrol and maladaptive perfectionism, cut across these subtypes. The clinically important question, then, may not be simply whether OCPD traits are present, but how they are organized within a given patient.

The organization and balance between traits may be critically important because different OCPD traits may disrupt PTSD treatment through different mechanisms. Emotional overcontrol is associated with internalizing symptoms, social isolation, and reduced emotional awareness (Hunt et al., 2022), as is the shame, self-criticism, and avoidance associated with maladaptive perfectionism. When these traits predominate, they may encumber effective PTSD treatment by reducing emotional disclosure, trust, flexible engagement, and distress tolerance, as well as limiting openness to practicing new behaviors. These patients may also struggle to describe their emotions, resist vulnerability, and may evaluate their own progress through harsh and inflexible standards. These traits may also intensify PTSD-related emotional dysregulation, contributing to pathological anger and over time, poorer social quality of life (Kulak et al., 2025).

Of special note, the combination of elevated perfectionism and emotional overcontrol has also been linked to elevated suicide risk (Hunt et al., 2022). In part, this risk may emerge from their direct interference with trauma-focused therapies. These patients may perseverate on treatment instructions, become preoccupied with doing exposure exercises “correctly,” or view treatment as ineffective unless it is conducted perfectly. They may argue with the rationale for exposure-based interventions, resist generalizing exposures to related contexts, or experience ordinary therapeutic uncertainty as evidence of failure. In this way, OCPD traits may not merely co-occur with PTSD symptoms, but may shape how PTSD is expressed, how treatment is received, and how clinical progress unfolds.

Unrecognized OCPD traits may also affect clinicians. Although empirical work in this area remains limited, civilian studies have linked OCPD with therapist disengagement, boredom, annoyance, and withdrawal (Colli et al., 2014). More broadly, negative countertransference has been linked to poorer therapeutic alliance and liklihood of treatment discontinuation across outpatient mental health clinics (Breivik Øvstebø et al., 2024). These dynamics are especially consequential in PTSD care, where clinician engagement and alliance are established predictors of outcome (Bauer et al., 2022; Sijercic et al., 2021). Although these findings come primarily from civilian samples, similar processes may be highly relevant in veteran care, as a healthcare system with fewer clinicians specializing in OCPD.

For these reasons, systematic screening for OCPD traits may improve care for a substantial subset of veterans receiving PTSD treatment. Brief self-report measures such as the POPS offer a practical way to identify clinically relevant traits (Pinto et al., 2022). Early identification may help providers anticipate treatment-interfering behaviors, tailor treatment plans, and frame rigidity, perfectionism, and emotional overcontrol as understandable but modifiable strategies that influence treatment response.

Treatment options for OCPD remain limited but are clinically actionable. No pharmacologic intervention is approved specifically for OCPD by the U.S. Food and Drug Association (FDA), though selective serotonin reuptake inhibitors may reduce maladaptive rigidity and perfectionism in some patients(Ansseau, 1994; Ekselius & von Knorring, 1998). In clinical practice, doses above the FDA label may be necessary, although empirical data supporting this remain limited. Specialized therapies such as cognitive behavioral therapy (CBT) and Acceptance and Commitment Therapy (ACT) (Cheli et al., 2020; Pinto, 2020; Pinto et al., 2022), radically-open dialectical behavioral therapy (Lynch et al., 2015, 2020; Steinhoff et al., 2026) and metacognitive therapy have shown efficacy for OCPD treatment (Gordon-King et al., 2018). When specialized therapy is unavailable, psychoeducation, brief CBT-based interventions, and bibliotherapy may be incorporated into medication management visits (Bowers et al., 2025). Early treatment efforts should focus on identifying rigidity and perfectionism as maladaptive, exploring how these behaviors are maintained. Behavioral experiments may be used to challenge harsh rules and standards, so long as they are designed to reinforce behavioral flexibility and self-compassion versus self-criticism. There are also numerous educational resources such as the book “Tightrope Walking” to support patients and families (Cheeseman, 2008). Skills training focused on emotion regulation and flexibility can also help psychosocial functioning (Cloitre et al., 2002).

This study has several important limitations. First, the sample was drawn from one psychiatrist’s outpatient practice within the VA medical system. Most patients in that caseload were transferred from other clinicians who had left the PTSD clinic, reducing the likelihood that the sample reflects systematic referral bias. However, the provider’s expertise in obsessive-compulsive and related disorders may have influenced more recent referrals, enriching the sample for OCPD. Despite this, we note that at 55.6%, prevalence of OCPD in this sample is only slightly higher than reported in other studies of veterans (range: 17-44%; Bollinger et al., 2000; Dunn et al., 2004; Southwick et al., 1993). Second, the sample was predominantly male and White, limiting generalizability across biological sex and race. Finally, the cross-sectional and retrospective design precludes conclusions about causality or the longitudinal impact of OCPD traits on PTSD course and treatment outcomes.

## Conclusion

In this chart review of outpatient veterans, OCPD traits were significantly associated with PTSD symptom severity, with emotional overcontrol and maladaptive perfectionism showing the strongest associations. These findings suggest that OCPD traits may be clinically important in Veterans with PTSD, not only as comorbid symptoms but as patterns that may shape distress, functioning, treatment engagement, and trauma recovery. Subsequent studies should examine how distinct configurations of OCPD traits influence PTSD presentation and outcomes, and whether systematic screening can improve case formulation, treatment planning, and long-term care for trauma-exposed Veterans.

## Data availability

The data that support the findings of this study may be available upon reasonable request to the corresponding author following all relevant regulations.

## Credit Authorship Contribution Statement

B.G. and J.B. conceived of the study and were responsible for study design. H.S. conducted statistical analyses. M.J.K., J.B. and T.Y.M. drafted the manuscript. T.S. and A.P. provided expert consultation and contributed to study design. All authors reviewed, edited, and approved the final manuscript.

## REFERENCES

Ansell, E. B., Pinto, A., Crosby, R. D., Becker, D. F., Añez, L. M., Paris, M., & Grilo, C. M. (2010). The prevalence and structure of obsessive-compulsive personality disorder in Hispanic psychiatric outpatients. Journal of Behavior Therapy and Experimental Psychiatry, 41(3), 275–281. 10.1016/j.jbtep.2010.02.005

Ansseau, M. (1994). Are SSRIs useful in obsessive-compulsive personality disorder? European Neuropsychopharmacology, 4(3), 266–267. 10.1016/0924-977X(94)90085-X

Bauer, A. G., Ruglass, L. M., Shevorykin, A., Saraiya, T. C., Robinson, G., Cadet, K., Julien, L., Chao, T., & Hien, D. (2022). Predictors of therapeutic alliance, treatment feedback, and clinical outcomes among African American women in treatment for co-occurring PTSD and SUD. Journal of Substance Abuse Treatment, 139, 108766. 10.1016/j.jsat.2022.108766

Beck, J. G., Grant, D. M., Clapp, J. D., & Palyo, S. A. (2009). Understanding the interpersonal impact of trauma: contributions of PTSD and depression. Journal of Anxiety Disorders, 23(4), 443–450. 10.1016/j.janxdis.2008.09.001

Bollinger, A. R., Riggs, D. S., Blake, D. D., & Ruzek, J. I. (2000). Prevalence of personality disorders among combat veterans with posttraumatic stress disorder. Journal of Traumatic Stress, 13(2), 255–270. 10.1023/A:1007706727869

Bowers, E. M., Levin, M. E., Ong, C. W., & Twohig, M. P. (2025). A randomized controlled trial of self-help acceptance and commitment therapy and cognitive behavioral therapy for perfectionism. Behaviour Research and Therapy, 192, 104806. 10.1016/j.brat.2025.104806

Breivik Øvstebø, R., Pedersen, G., Wilberg, T., Røssberg, J. I., Johnsen Dahl, H.-S., & Kvarstein, E. H. (2024). Countertransference, alliance, and outcome in the treatment of patients with personality disorder: a longitudinal naturalistic study. Frontiers in Psychiatry, 15. 10.3389/fpsyt.2024.1490056

Campbell-Sills, L., Sun, X., Choi, K. W., He, F., Ursano, R. J., Kessler, R. C., Levey, D. F., Smoller, J. W., Gelernter, J., Jain, S., & Stein, M. B. (2022). Dissecting the heterogeneity of posttraumatic stress disorder: differences in polygenic risk, stress exposures, and course of PTSD subtypes. Psychological Medicine, 52(15), 3646–3654. 10.1017/S0033291721000428

Cheeseman, G. D. (2008). Tightrope Walking: All You Need to Know about OCPD and Perfectionism. Willows Brooks Publishing.

Cheli, S., MacBeth, A., Popolo, R., & Dimaggio, G. (2020). The intertwined path of perfectionism and self-criticism in a client with obsessive-compulsive personality disorder. Journal of Clinical Psychology, 76(11), 2055–2066. 10.1002/jclp.23051

Clemente, M. J., Martins Silva, A. S., Pozzolo Pedro, M. O., Paiva, H. S., de Azevedo Marques Périco, C., Torales, J., Ventriglio, A., & Castaldelli-Maia, J. M. (2022). A meta-analysis and meta-regression analysis of the global prevalence of obsessive-compulsive personality disorder. Heliyon, 8(7), e09912. 10.1016/j.heliyon.2022.e09912

Cloitre, M., Koenen, K. C., Cohen, L. R., & Han, H. (2002). Skills training in affective and interpersonal regulation followed by exposure: A phase-based treatment for PTSD related to childhood abuse. Journal of Consulting and Clinical Psychology, 70(5), 1067–1074.

Colli, A., Tanzilli, A., Dimaggio, G., & Lingiardi, V. (2014). Patient Personality and Therapist Response: An Empirical Investigation. American Journal of Psychiatry, 171(1), 102–108. 10.1176/appi.ajp.2013.13020224

Diagnostic and Statistical Manual of Mental Disorders (5th ed.). (2022). American Psychiatric Association.

Dunn, N. J., Yanasak, E., Schillaci, J., Simotas, S., Rehm, L. P., Souchek, J., Menke, T., Ashton, C., – Hamilton, J. D. (2004). Personality disorders in veterans with posttraumatic stress disorder and depression. Journal of Traumatic Stress, 17(1), 75–82. 10.1023/B:JOTS.0000014680.54051.50

Edwards, E. R., Tran, H., Wrobleski, J., Rabhan, Y., Yin, J., Chiodi, C., Goodman, M., & Geraci, J. (2022). Prevalence of Personality Disorders Across Veteran Samples: A Meta-Analysis. Journal of Personality Disorders, 36(3), 339–358. 10.1521/pedi.2022.36.3.339

Ekselius, L., & von Knorring, L. (1998). Personality disorder comorbidity with major depression and response to treatment with sertraline or citalopram. International Clinical Psychopharmacology, 13(5), 205–212. 10.1097/00004850-199809000-00003

Gerdan, G., & Salcioglu, E. (2025). Experiential Avoidance as a Transdiagnostic Mediator in the Relationship Between Intolerance of Uncertainty, Maladaptive Perfectionism, and Psychiatric Symptoms: Structural and Causal Mediation Analyses in a Clinical Sample. Clinical Psychology & Psychotherapy, 32(3). 10.1002/cpp.70102

Goodman, W. K., Price, L. H., Rasmussen, S. A., Mazure, C., Delgado, P., Heninger, G. R., & Charney, D. S. (1989). The Yale-Brown Obsessive Compulsive Scale. II. Validity. Archives of General Psychiatry, 46(11), 1012–1016. 10.1001/archpsyc.1989.01810110054008

Goodman, W. K., Price, L. H., Rasmussen, S. A., Mazure, C., Fleischmann, R. L., Hill, C. L., Heninger, G. R., & Charney, D. S. (1989). The Yale-Brown Obsessive Compulsive Scale. I. Development, use, and reliability. Archives of General Psychiatry, 46(11), 1006–1011. 10.1001/archpsyc.1989.01810110048007

Gordon-King, K., Schweitzer, R. D., & Dimaggio, G. (2018). Metacognitive Interpersonal Therapy for Personality Disorders Featuring Emotional Inhibition. Journal of Nervous & Mental Disease, 206(4), 263–269. 10.1097/NMD.0000000000000789

Grant, J. E., Mooney, M. E., & Kushner, M. G. (2012). Prevalence, correlates, and comorbidity of DSM-IV obsessive-compulsive personality disorder: Results from the National Epidemiologic Survey on Alcohol and Related Conditions. Journal of Psychiatric Research, 46(4), 469–475. 10.1016/j.jpsychires.2012.01.009

Grau, P. P., Sripada, R. K., Pietrzak, R. H., Ganoczy, D., & Harpaz-Rotem, I. (2022). Treatment response trajectories in residential PTSD programs for veterans: A national cohort investigation. Journal of Anxiety Disorders, 92, 102645. 10.1016/j.janxdis.2022.102645

Hunt, C., Exline, J. J., Fletcher, T. L., & Teng, E. J. (2022). Intolerance of uncertainty prospectively predicts the transdiagnostic severity of emotional psychopathology: Evidence from a Veteran sample. Journal of Anxiety Disorders, 86, 102530. 10.1016/j.janxdis.2022.102530

Kelly, M. M., DeBeer, B. B., Meyer, E. C., Kimbrel, N. A., Gulliver, S. B., & Morissette, S. B. (2019). Experiential avoidance as a mediator of the association between posttraumatic stress disorder symptoms and social support: A longitudinal analysis. Psychological Trauma : Theory, Research, Practice and Policy, 11(3), 353–359. 10.1037/tra0000375

Kulak, M. J., Gonsalves, M. A., Mariano, T. Y., Pinto, A., Greenberg, B. D., Barredo, J., Kunicki, Z., & Shea, M. T. (2025). An exploratory analysis of obsessive-compulsive personality traits, pathologic anger and quality of life among trauma-exposed veterans. Comprehensive Psychiatry, 143, 152628. 10.1016/j.comppsych.2025.152628

Lynch, T. R., Hempel, R. J., Whalley, B., Byford, S., Chamba, R., Clarke, P., Clarke, S., Kingdon, D. G., O’Mahen, H., Remington, B., Rushbrook, S. C., Shearer, J., Stanton, M., Swales, M., Watkins, A., & Russell, I. T. (2020). Refractory depression – mechanisms and efficacy of radically open dialectical behaviour therapy (RefraMED): findings of a randomised trial on benefits and harms. The British Journal of Psychiatry, 216(4), 204–212. 10.1192/bjp.2019.53

Lynch, T. R., Whalley, B., Hempel, R. J., Byford, S., Clarke, P., Clarke, S., Kingdon, D., O’Mahen, H., Russell, I. T., Shearer, J., Stanton, M., Swales, M., Watkins, A., & Remington, B. (2015). Refractory depression: mechanisms and evaluation of radically open dialectical behaviour therapy (RO-DBT) [REFRAMED]: protocol for randomised trial. BMJ Open, 5(7), e008857. 10.1136/bmjopen-2015-008857

McLean, C. P., & Foa, E. B. (2017). Emotions and emotion regulation in posttraumatic stress disorder. Current Opinion in Psychology, 14, 72–77. 10.1016/j.copsyc.2016.10.006

Patel, T. A., Blakey, S. M., Halverson, T. F., Mann, A. J. D., Calhoun, P. S., Beckham, J. C., Pugh, M. J., & Kimbrel, N. A. (2023). Experiential Avoidance, Posttraumatic Stress Disorder, and Self-Injurious Thoughts and Behaviors: A Moderation Analysis in a National Veteran Sample. International Journal of Cognitive Therapy, 1. 10.1007/s41811-023-00164-2

Pinto, A. (2020). Psychotherapy for obsessive compulsive personality disorder. In J. E. Grant, A. Pinto, & S. Chamberlain (Eds.), Obsessive Compulsive Personality Disorder. American Psychiatric Association Publishing.

Pinto, A., Ansell, E. B., & Wright, A. G. C. (2011). A new approach to the assessment of obsessive compulsive personality. Annual Meeting of the Society for Personality Assessment.

Pinto, A., Teller, J., & Wheaton, M. G. (2022). Obsessive-Compulsive Personality Disorder: A Review of Symptomatology, Impact on Functioning, and Treatment. Focus (American Psychiatric Publishing), 20(4), 389–396. 10.1176/appi.focus.20220058

Rassu, F. S., Sansgiry, S., Hundt, N. E., Kunik, M. E., & Cully, J. A. (2022). Presence of PTSD is Associated with Clinical and Functional Impact in Veterans with Depression Treated in Community-Based Clinics. Journal of Clinical Psychology in Medical Settings, 29(1), 220–229. 10.1007/s10880-021-09796-y

Riddle, M. A., Maher, B. S., Wang, Y., Grados, M., Bienvenu, O. J., Goes, F. S., Cullen, B., Murphy, D. L., Rauch, S. L., Greenberg, B. D., Knowles, J. A., McCracken, J. T., Pinto, A., Piacentini, J., Pauls, D. L., Rasmussen, S. A., Shugart, Y. Y., Nestadt, G., & Samuels, J. (2016). OBSESSIVE-COMPULSIVE PERSONALITY DISORDER: EVIDENCE FOR TWO DIMENSIONS. Depression and Anxiety, 33(2), 128–135. 10.1002/da.22452

Sadri, S. K., McEvoy, P. M., Pinto, A., Anderson, R. A., & Egan, S. J. (2019). A Psychometric Examination of the Pathological Obsessive Compulsive Personality Scale (POPS): Initial Study in an Undergraduate Sample. Journal of Personality Assessment, 101(3), 284–293. 10.1080/00223891.2018.1428983

Sijercic, I., Liebman, R. E., Stirman, S. W., & Monson, C. M. (2021). The Effect of Therapeutic Alliance on Dropout in Cognitive Processing Therapy for Posttraumatic Stress Disorder. Journal of Traumatic Stress, 34(4), 819–828. 10.1002/jts.22676

Southwick, S. M., Yehuda, R., & Giller, E. L. (1993). Personality disorders in treatment-seeking combat veterans with posttraumatic stress disorder. The American Journal of Psychiatry, 150(7), 1020–1023. 10.1176/ajp.150.7.1020

Steinhoff, M. F., Baudinet, J., Hempel, R. J., Tillman, R., Lynch, T. R., & Gilbert, K. E. (2026). Obsessive-compulsive personality disorder in radically open dialectical behavior therapy for treatment-refractory depression. Personality Disorders, 17(2), 178–188. 10.1037/per0000754

Teller, J. A., Ward, H., Jennings, A. F., Arcaro, N., Christman, J., Wheaton, M. G., & Pinto, A. (2026). Examining the association between OCPD and OCD: Data from a specialized outpatient clinic. Journal of Obsessive-Compulsive and Related Disorders, 48, 100993. 10.1016/j.jocrd.2025.100993

Weathers, F. (2013). The PTSD Checklist for the DSM-5 (PCL-5).

Weathers, F. W., Bovin, M. J., Lee, D. J., Sloan, D. M., Schnurr, P. P., Kaloupek, D. G., Keane, T. M., & Marx, B. P. (2018). The Clinician-Administered PTSD Scale for DSM-5 (CAPS-5): Development and initial psychometric evaluation in military veterans. Psychological Assessment, 30(3), 383–395. 10.1037/pas0000486

